## Supplementary figures and images for "Shedding of Infectious SARS-CoV-2 Despite Vaccination"

### Supplemental Figure 1

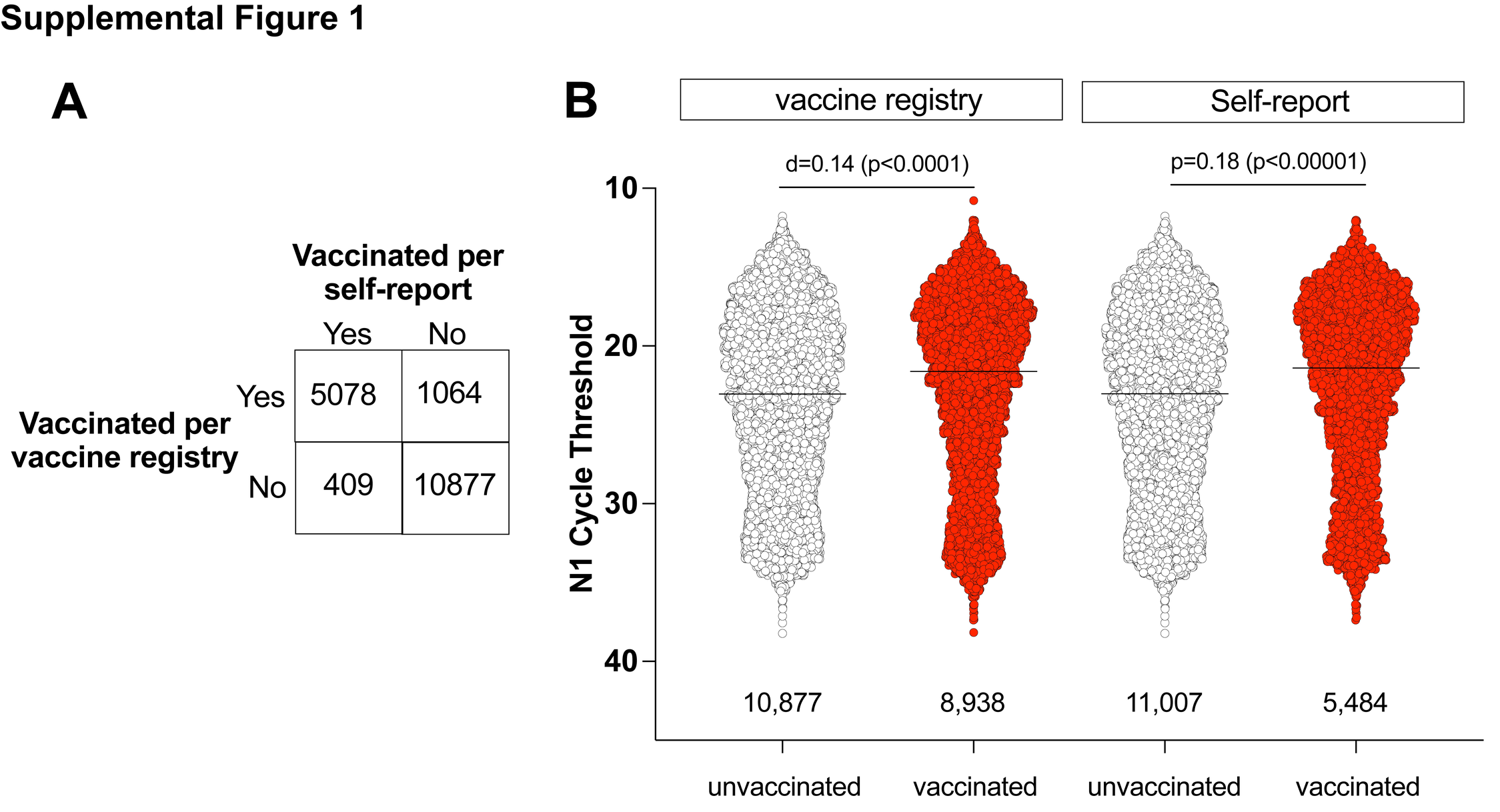

### Supplemental Figure 2

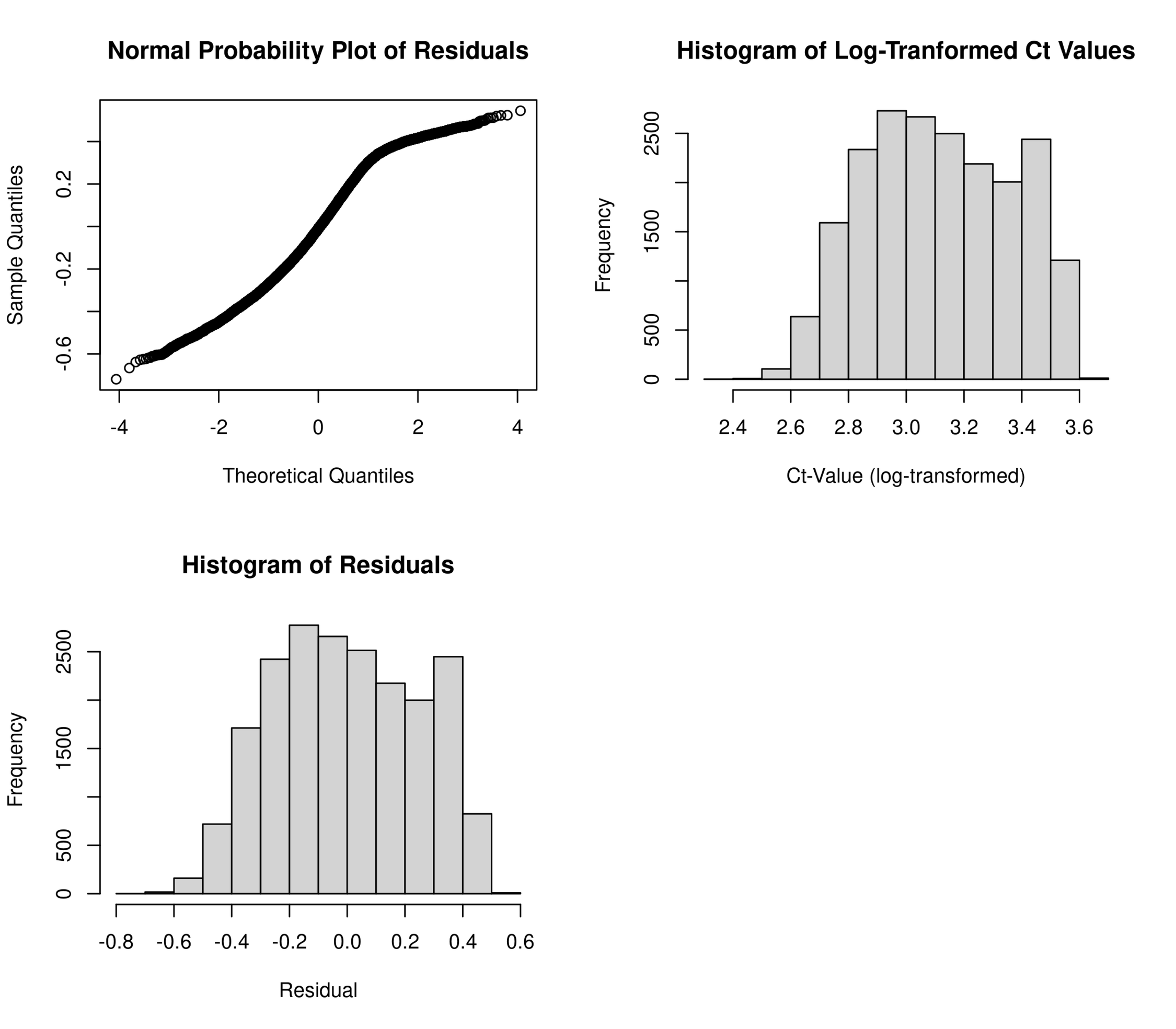

### Supplemental Figure 3

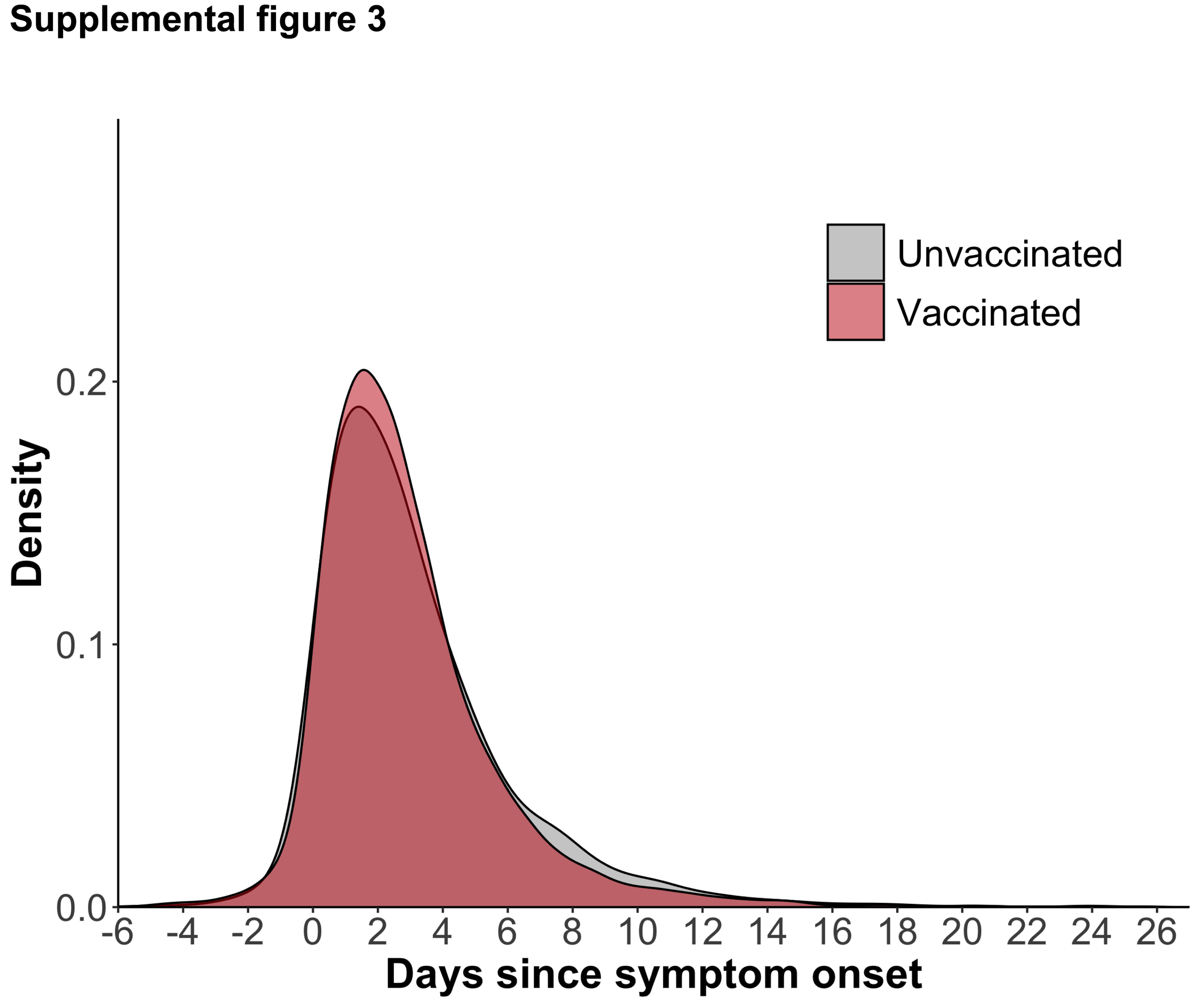

### Supplemental Figure 4

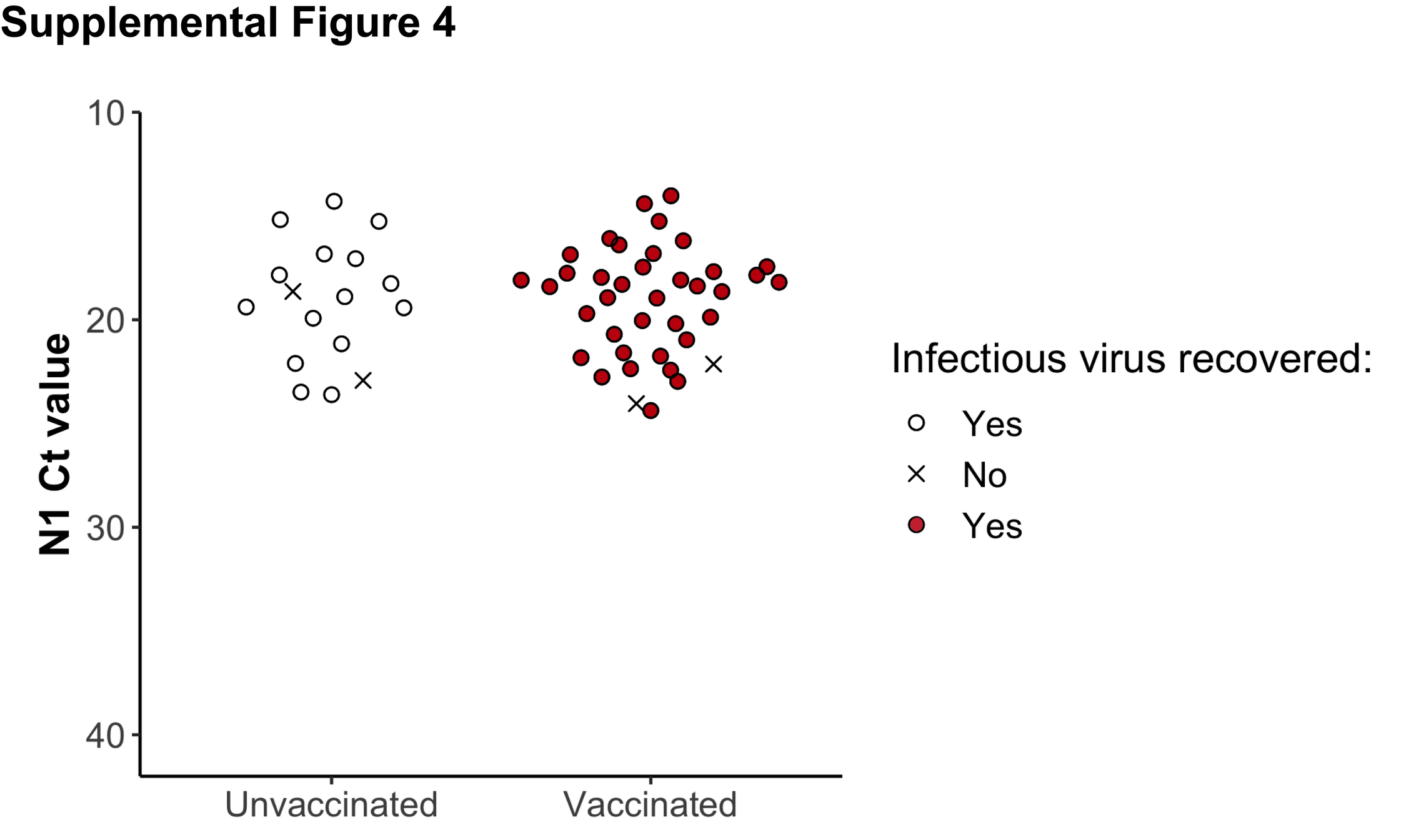

### Supplemental Figure 5

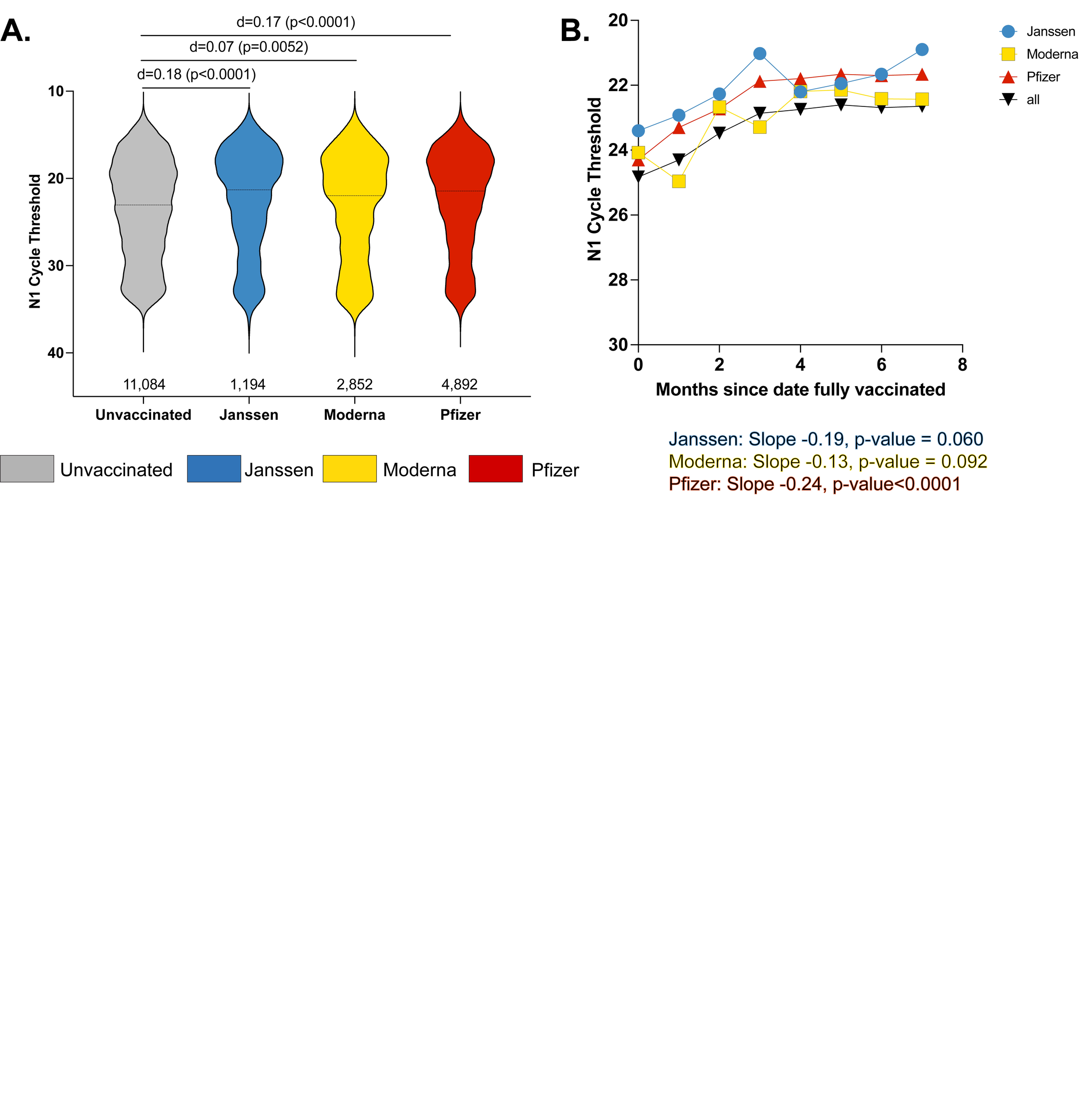
