## Supplemental Tables and Figure legends for "Shedding of Infectious SARS-CoV-2 Despite Vaccination"

**Supplemental Table 1**: *Comparisons between vaccine type*

|  | **Mean** | **95% CI** |
| --- | --- | --- |
| Unvaccinated | 22.9 | 22.8-23.0 |
| Janssen | 21.9 | 21.6-22.2 |
| Moderna | 22.5 | 22.3-22.7 |
| Pfizer | 22.0 | 21.8-22.1 |

| **p-** | **p-** | **p-** | **p-** | **p-** | **p-** | **p-** |
| --- | --- | --- | --- | --- | --- | --- |
| **value±** | **value^1^** | **value^2^** | **value^3^** | **value^4^** | **value^5^** | **value^6^** |
| <0.0001 | <0.0001 | 0.0052 | <0.0001 | 0.0064 | 0.9870 | 0.0001 |

‡ comparisons between all groups

**^1^**: comparison Unvaccinated vs. Janssen (adjusted for multiple comparisons using Tukey’s HSD method) **^2^**: comparison Unvaccinated vs. Moderna (adjusted for multiple comparisons using Tukey’s HSD method) **^3^**: comparison Unvaccinated vs. Pfizer (adjusted for multiple comparisons using Tukey’s HSD method)

**^4^**: comparison Janssen vs. Moderna (adjusted for multiple comparisons using Tukey’s HSD method)

**^5^**: comparison Janssen vs. Pfizer (adjusted for multiple comparisons using Tukey’s HSD method)

**^6^**: comparison Moderna vs. Pfizer (adjusted for multiple comparisons using Tukey’s HSD method)

**Supplemental Table 2**: *Comparison of Ct values in vaccinated and unvaccinated persons, stratified by age group (there is a significant interaction between age group and vaccination status, p<0.0001)*

|  | **Not Vaccinated**  **Mean 95% CI** | | **Vacci**  **Mean** | **nated**  **95% CI** | **Effect size *d*** | **p-value** |
| --- | --- | --- | --- | --- | --- | --- |
| 0-11 yr | 23.9 | 23.7-24.1 | 19.8 | 16.5-23.8 | 0.79 | 0.0466 |
| 12-18 yr | 23.0 | 22.8-23.3 | 23.9 | 22.5-23.5 | 0.00 | 0.9242 |
| 19-35 yr | 22.4 | 22.2-22.6 | 23.0 | 22.1-22.6 | 0.00 | 0.8846 |
| 36-60 yr | 22.3 | 22.1-22.5 | 21.9 | 21.8-22.1 | 0.07 | 0.0080 |
| >61 yr | 22.3 | 21.9-22.8 | 22.1 | 21.8-22.3 | 0.05 | 0.3239 |

**Supplemental Table 3**: *Comparison of Ct values in vaccinated and unvaccinated persons, stratified by sex.*

|  | **Unvaccinated**  **Mean 95% CI** | **Vaccinated**  **Mean 95% CI** | **Effect size *d*** | **p-value** |
| --- | --- | --- | --- | --- |
| Female | 23.0 22.9-23.2 | 22.3 22.1-22.4 | 0.14 | <0.0001 |
| Male | 22.8 22.6-22.9 | 22.0 21.8-22.1 | 0.15 | <0.0001 |

**Supplemental Figure 1. Concordance between self-reported vaccination status and records in public health vaccine registries.** Individuals were considered fully vaccinated based on vaccine registry (WIR/WEDSS) data if the registries indicated receipt of a final vaccine dose at least 14 days prior to submitting the sample used in our analysis. For individuals whose vaccination status could not be verified in the registry, self-reported data collected at the time of testing were used. Individuals were considered unvaccinated based on self-report only if there was an explicit declaration of unvaccinated status in the self-reported data. Individuals were considered fully vaccinated based on self-report if they fulfilled all of the following criteria: (1) indicated that they had received a COVID vaccine prior to testing; (2) indicated that they did not require another vaccine dose; and (3) reported a date of last vaccine dose that was at least 14 days prior to testing.
